# Unlocking the Treatment of Facioscapulohumeral Muscular Dystrophy Type 2: The Bisphenol Connection

**DOI:** 10.1101/2024.03.12.24304159

**Authors:** Saed Sayad, Mark Hiatt, Hazem Mustafa

## Abstract

**Background:** Facioscapulohumeral muscular dystrophy type 2 (FSHD2) poses a significant challenge within the domain of neuromuscular disorders, marked by a progressive decline in muscle strength accompanied by tissue wasting. FSHD2 results from chromosomal deletions triggering the activation of a dormant gene known as DUX4. While DUX4 typically regulates early embryonic development, its activation in adult muscle cells leads to premature cell death. Despite this understanding, the exact pathology of FSHD2 remains unclear. To date, no effective treatment for FSHD2 exists.

**Method:** We acquired single-cell RNA sequencing (RNA-Seq) data (*GSE143452*) from primary myoblasts for FSHD2 from the United States National Institutes of Health (NIH) portal website. Our analysis encompassed a comprehensive examination of differentially expressed genes, alongside associated compounds sourced from the Chemical Entities of Biological Interest ([ChEBI](https://www.ebi.ac.uk/chebi/)) database. Employing rigorous statistical methods, we pinpointed the most prominently upregulated and downregulated genes. Subsequently, we determined the compounds capable of modulating the expression of these top genes, either enhancing or reducing their activity.

**Results:** Bisphenol S (BPS) can upregulate 52 of 100 top downregulated genes in FSHD2 without downregulating any other genes and Bisphenol F (BPF) can upregulate 45 of 100 downregulated genes with downregulating only one other gene. The enrichment analysis of both sets of 52 genes related to BPS and 45 genes corresponding to BPF highlights their significant involvement in various aspects of muscle biology, particularly as pertaining to the function and dysfunction of cardiac and skeletal muscle.

**Conclusions:** Leveraging single-cell RNA-Seq data and computational analysis, we identified key dysregulated genes in FSHD2 and elucidated their modulation by compounds such as BPS and BPF. While effective treatments for FSHD2 remain elusive, our study provides valuable insights into potential therapeutic targets and pathways for further investigation in the pursuit of effective interventions for this debilitating condition. However, more research is needed to understand whether the roles of BPS and F are constructive or destructive.

## Introduction

Facioscapulohumeral muscular dystrophy (FSHD) is a genetic condition marked by a gradual weakening of muscles, primarily in the face, shoulders, and upper arms [1]. There are two types: FSHD type 1, linked to a deletion on chromosome 4 (D4Z4), and FSHD type 2 (FSHD2), caused by mutations in the SMCHD1 gene. Both result in the activation of a dormant gene called DUX4, which normally plays a role in embryonic development but, when activated in adult muscle cells, leads to premature cell death. Despite advancements, the exact mechanisms behind FSHD2 remain incompletely understood. Currently, no cure for FSHD2 exists, with treatment focusing on managing symptoms and improving the quality of life. Research into potential targeted therapies and gene-based treatments for FSHD2 is ongoing, offering hope for future breakthroughs in disease management and ultimately a cure. Single-cell transcriptomics offer a powerful tool to investigate the molecular processes and gene-expression changes associated with FSHD2, particularly focusing on differentially expressed genes and conducting pathway, gene ontology, and gene-compound analyses within the context of FSHD2.

## Data

We acquired single-cell transcriptomes (*GSE143452*) from primary myoblasts for FSHD2 from the United States National Institutes of Health (NIH) portal website (**Figure 1**). All gene-related data have been sourced from the [GeneCards](https://www.genecards.org/) website. We used the [g:Profiler](https://biit.cs.ut.ee/gprofiler/gost) web service to conduct enrichment analysis, leveraging its powerful tools to elucidate gene function. Additionally, we utilized the Chemical Entities of Biological Interest ([ChEBI](https://www.ebi.ac.uk/chebi/)) database to explore gene-compound relationships, drawing insights from its comprehensive collection of chemical entities of biological relevance.

**Figure 1:**
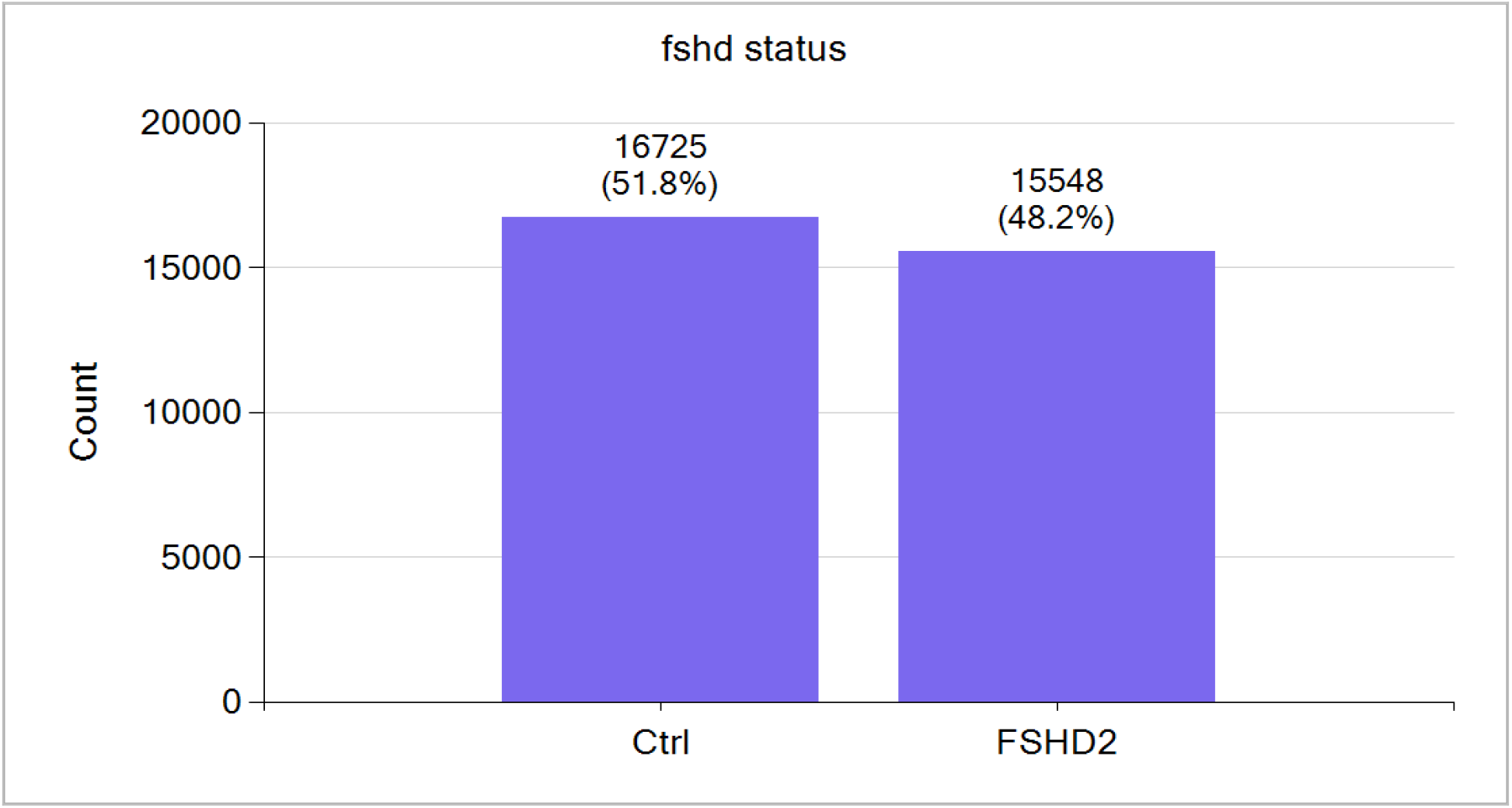
*GSE143452* comprises 16,725 control single cells and 15,548 FSHD2 single cells. The total number of genes is 19,615.

## Data Analysis

Employing rigorous statistical techniques, we identified the differentially expressed genes displaying significant upregulation and downregulation. Furthermore, we delved deeper to discern compounds capable of modulating the expression of these critical genes, shedding light on potential avenues for therapeutic intervention or further experimental inquiry. Our analysis encompassed a comprehensive examination of differentially expressed genes, alongside associated compounds sourced from the ChEBI database. Again employing precise statistical methods, we pinpointed the most prominently upregulated and downregulated genes. Subsequently, we determined the compounds capable of modulating the expression of these top genes, either enhancing or reducing their activity.

## Upregulated and Downregulated Differentially Expressed Genes

The single-cell gene expression analysis (**Table 1**) has revealed the top 10 upregulated genes. TIMP1 is known for its role in regulating metalloproteinase activity and the extracellular matrix [ 2]. IGFBP7 is associated with pathways involving insulin-like growth factors and cell growth regulation [3]. FN1 participates in various cellular processes such as cell adhesion and migration, crucial in embryogenesis, wound healing, blood coagulation, host defense, and metastasis [4]. CCN2, secreted by vascular endothelial cells, acts as a mitogen influencing chondrocyte proliferation and differentiation, as well as cell adhesion in diverse cell types [5]. CTSB may initiate proteolytic pathways important for inflammatory breast cancer invasion and is considered a potential prognostic marker for lymphatic metastasis [6]. TMSB4X encodes an actin-sequestering protein involved in regulating actin polymerization, cell proliferation, migration, and differentiation. IFITM3 plays a role in regulating fibrinogen endocytosis and platelet reactivity in nonviral sepsis. MT2A’s function spans metal homeostasis regulation, detoxification, oxidative stress management, immune defense, cell-cycle progression, cell proliferation and differentiation, and angiogenesis [7]. ACTG1 is essential for muscle contraction, and any alterations in actin expression could directly affect muscle function and integrity [8]. SERPINE2, encoding a member of the serpin family, acts as an inhibitor of serine proteases [9].

**Table 1:** Top 10 upregulated and downregulated differentially expressed genes through single-cell RNA-Seq.

| Top 10 Upregulated and Downregulated Differentially Expressed Genes |  |  |  |
| --- | --- | --- | --- |
| Upregulated Genes |  | Downregulated Genes |  |
| Gene Symbol | Gene Name | Gene Symbol | Gene Name |
| *TIMP1* | Metallopeptidase Inhibitor 1 | *ACTC1* | Actin Alpha Cardiac Muscle 1 |
| *IGFBP7* | *Insulin Like Growth Factor Binding Protein 7* | *ACTA1* | Actin Alpha 1, Skeletal Muscle |
| *FN1* | Fibronectin 1 | *MYL11* | Myosin Light Chain 11 |
| *CCN2* | Cellular Communication Network Factor 2 | *MYH3* | Myosin Heavy Chain 3 |
| *CTSB* | Cathepsin B | *MYL6B* | Myosin Light Chain 6B |
| *TMSB4X* | Thymosin Beta 4 X-Linked | *MYBPH* | Myosin Binding Protein H |
| *IFITM3* | Interferon Induced Transmembrane Protein 3 | *TPM2* | Tropomyosin 2 |
| *MT2A* | Metallothionein 2A | *MYL2* | Myosin Light Chain 2 |
| *ACTG1* | Actin Gamma 1 | *MYL1* | Myosin Light Chain 1 |
| *SERPINE2* | Serpin Family E Member 2 | *TNNI1* | Troponin I1, Slow Skeletal Type |

The single-cell analysis of FSHD2 has unveiled a significant downregulation of crucial genes essential for muscle function. Among the top 10 downregulated genes are ACTC1, ACTA1, MYL11, MYH3, MYL6B, MYBPH, TPM2, MYL2, MYL1, and TNNI1, each playing pivotal roles in muscle contraction and skeletal muscle integrity. These genes encode proteins integral to the structural organization and dynamic regulation of the sarcomere, the basic contractile unit of muscle fibers. For instance, ACTC1 and ACTA1 encode actin proteins, which form the thin filaments of the sarcomere, while MYH3 encodes a myosin protein crucial for muscle contraction. Downregulation of these genes suggests a potential disruption in contractility, which could have profound implications for muscle function and integrity in FSHD2. Understanding the regulatory mechanisms governing these genes holds promise for advancing our knowledge of FSHD2 pathogenesis and developing targeted interventions aimed at restoring muscle function [10].

## Gene-Compound Analysis

We mined the ChEBI database to identify compounds with similar or contrasting regulatory patterns to the top 10 upregulated and downregulated genes. Notably, Bisphenol F (BPF) and Bisphenol S (BPS) were found to upregulate eight and seven, respectively, of the top 10 downregulated genes, while not affecting any of the top 10 upregulated genes. Our investigation extended to the top 100 downregulated genes, revealing that BPS upregulate 52 genes without downregulating any others. **Table 2** shows 52 genes from the top 100 downregulated list, which BPS can upregulate.

**Table 2:** 52 of the top 100 downregulated genes that can be upregulated by BPS.

|  |  |  |  |  |  |  |  |  |  |  |
| --- | --- | --- | --- | --- | --- | --- | --- | --- | --- | --- |
| ACTC1 | MYL1 | ENO3 | CASQ2 | H19 | ITGA7 | HSPB1 | NEB | EHBP1L1 | MYL3 | COX7A1 |
| MYLPF | TNNI1 | DES | MYH8 | KLHL41 | ALDOA | PRUNE2 | LIMCH1 | MYOG | COX6A2 | ELN |
| MYH3 | CRYAB | ANKRD1 | DCN | SYNPO2 | TNNI2 | HTRA1 | HRC | MYOM2 | SSPN |  |
| TPM2 | MYH7 | TPM1 | HSPB8 | MEF2C | FLNC | LUM | ALDH1A1 | MYOM1 | PGAM2 |  |
| MYL2 | TTN | TNNC2 | ACTN2 | NES | TNNC1 | CSRP3 | SYNPO2L | GYS1 | NEXN |  |

Furthermore, BPF upregulates 45 genes from this list, with only one exception in which it downregulates a single gene (HSPB2) among the top 100 downregulated genes. **Table 3** showcases 45 genes from the same list upregulated by BPF.

**Table 3:** 45 of the top 100 downregulated genes that can be upregulated by BPF.

|  |  |  |  |  |  |  |  |  |
| --- | --- | --- | --- | --- | --- | --- | --- | --- |
| ACTC1 | MYL2 | TNNT3 | ANKRD1 | MYH8 | KLHL41 | TNNC1 | LIMCH1 | MYOG |
| ACTA1 | MYL1 | TTN | TPM1 | DCN | SYNPO2 | HTRA1 | HRC | MYOM2 |
| MYLPF | TNNI1 | ENO3 | TNNC2 | HSPB8 | MEF2C | LUM | ALDH1A1 | MYL3 |
| MYL6B | CRYAB | DES | TNNT1 | ACTN2 | NES | CSRP3 | SYNPO2L | PGAM2 |
| TPM2 | MYH7 | TNNT2 | CASQ2 | H19 | TNNI2 | NEB | EHBP1L1 | ELN |

## Functional-Enrichment Analysis

In the next step, we used [g:Profiler](https://biit.cs.ut.ee/gprofiler/gost), a web server designed for functional-enrichment analysis and conversions of gene lists. As a means to gain insights into the biological functions, pathways, and processes associated with a given set of genes or proteins, this tool is widely used in bioinformatics and systems biology research to interpret high-throughput experimental data and understand their underlying biological mechanisms.

In **Table 4**, the enrichment analysis of 52 genes from Table 2 using g:Profiler reveals a comprehensive involvement in muscle biology and cardiac function. The genes are notably associated with various aspects of muscle physiology, including contraction, development, and structural organization, as evidenced by enriched terms in gene ontology categories such as muscle system process and myofibril assembly. Additionally, pathways related to cardiac muscle contraction and cardiomyopathy, as well as cytoskeletal protein binding and metabolic processes, are significantly enriched. Human phenotype ontology analysis highlights associations with clinical manifestations like abnormal cardiac function and skeletal muscle abnormalities. These findings underscore the genetic underpinnings of muscular and cardiac physiology, providing insights into potential mechanisms underlying muscle-related disorders and cardiomyopathies.

**Table 4:** Functional enrichment analysis of 52 genes upregulated by BPS (listed in Table 2) using g:Profiler.

| Source | Name | Native |
| --- | --- | --- |
| CORUM | MYOM1 homodimer complex | CORUM:1987 |
| GO:BP | muscle contraction | GO:0006936 |
| GO:BP | muscle system process | GO:0003012 |
| GO:BP | muscle organ development | GO:0007517 |
| GO:BP | muscle structure development | GO:0061061 |
| GO:BP | myofibril assembly | GO:0030239 |
| GO:CC | contractile fiber | GO:0043292 |
| GO:CC | sarcomere | GO:0030017 |
| GO:CC | myofibril | GO:0030016 |
| GO:CC | supramolecular fiber | GO:0099512 |
| GO:CC | supramolecular polymer | GO:0099081 |
| GO:MF | structural constituent of muscle | GO:0008307 |
| GO:MF | actin binding | GO:0003779 |
| GO:MF | cytoskeletal protein binding | GO:0008092 |
| GO:MF | actin filament binding | GO:0051015 |
| GO:MF | structural molecule activity | GO:0005198 |
| HP | EMG abnormality | HP:0003457 |
| HP | Orthopnea | HP:0012764 |
| HP | Left ventricular systolic dysfunction | HP:0025169 |
| HP | Abnormal left ventricular function | HP:0005162 |
| HP | Exertional dyspnea | HP:0002875 |
| HPA | skeletal muscle; myocytes[High] | HPA:0440343 |
| HPA | skeletal muscle; myocytes[≥Medium] | HPA:0440342 |
| HPA | heart muscle; cardiomyocytes[High] | HPA:0241103 |
| HPA | skeletal muscle | HPA:0440000 |
| HPA | skeletal muscle; myocytes[≥Low] | HPA:0440341 |
| KEGG | Motor proteins | KEGG:04814 |
| KEGG | Cardiac muscle contraction | KEGG:04260 |
| KEGG | Hypertrophic cardiomyopathy | KEGG:05410 |
| KEGG | Dilated cardiomyopathy | KEGG:05414 |
| KEGG | Adrenergic signaling in cardiomyocytes | KEGG:04261 |
| REAC | Striated Muscle Contraction | REAC:R-has-390522 |
| REAC | Muscle contraction | REAC:R-has-397014 |
| REAC | Metabolism of carbohydrates | Rhas:R-HSA-71387 |
| WP | Striated muscle contraction pathway | WP:WP383 |
| WP | Cardiomyocyte signaling pathways converging on Titin | WP:WP5344 |

In **Table 5**, analysis of the 45 genes from Table 3 highlights the significant involvement of these input genes in various aspects of muscle biology, particularly as pertaining to cardiac and skeletal muscle function. These genes are associated with processes such as muscle contraction and structure development and myofibril assembly, along with molecular functions like binding of cytoskeletal protein and actin. Phenotypic associations include such electromyographic (EMG) and cardiac abnormalities as left ventricular systolic dysfunction and hypertrophic cardiomyopathy. Enriched pathways encompass striated muscle contraction, myocardial signaling, and pathways related to cardiomyocyte differentiation and function. Overall, these findings emphasize the crucial roles of the input genes in muscle physiology and pathology, particularly in cardiac and skeletal muscle systems.

**Table 5:** The functional-enrichment analysis of 45 genes upregulated by BPF (listed in Table 3) using g:Profiler.

| Source | Name | Native |
| --- | --- | --- |
| GO:BP | muscle contraction | GO:0006936 |
| GO:BP | muscle system process | GO:0003012 |
| GO:BP | myofibril assembly | GO:0030239 |
| GO:BP | striated muscle cell development | GO:0055002 |
| GO:BP | muscle structure development | GO:0061061 |
| GO:CC | contractile fiber | GO:0043292 |
| GO:CC | sarcomere | GO:0030017 |
| GO:CC | myofibril | GO:0030016 |
| GO:CC | supramolecular fiber | GO:0099512 |
| GO:CC | supramolecular polymer | GO:0099081 |
| GO:CC | supramolecular complex | GO:0099080 |
| GO:MF | structural constituent of muscle | GO:0008307 |
| GO:MF | cytoskeletal protein binding | GO:0008092 |
| GO:MF | actin binding | GO:0003779 |
| GO:MF | structural molecule activity | GO:0005198 |
| GO:MF | myosin binding | GO:0017022 |
| HP | EMG abnormality | HP:0003457 |
| HP | Left ventricular systolic dysfunction | HP:0025169 |
| HP | Exertional dyspnea | HP:0002875 |
| HP | Myopathy | HP:0003198 |
| HP | Orthopnea | HP:0012764 |
| HPA | skeletal muscle; myocytes[High] | HPA:0440343 |
| HPA | heart muscle; cardiomyocytes[High] | HPA:0241103 |
| HPA | skeletal muscle; myocytes[≥Medium] | HPA:0440342 |
| HPA | skeletal muscle; myocytes[≥Low] | HPA:0440341 |
| HPA | skeletal muscle | HPA:0440000 |
| KEGG | Motor proteins | KEGG:04814 |
| KEGG | Cardiac muscle contraction | KEGG:04260 |
| KEGG | Hypertrophic cardiomyopathy | KEGG:05410 |
| KEGG | Dilated cardiomyopathy | KEGG:05414 |
| KEGG | Adrenergic signaling in cardiomyocytes | KEGG:04261 |
| REAC | Striated Muscle Contraction | REAC:R-HSA-390522 |
| REAC | Muscle contraction | REAC:R-HSA-397014 |
| REAC | Smooth Muscle Contraction | REAC:R-HSA-445355 |
| WP | Striated muscle contraction pathway | WP:WP383 |
| WP | Cardiomyocyte signaling pathways converging on Titin | WP:WP5344 |
| WP | Cardiac progenitor differentiation | WP:WP2406 |

## Bisphenol S

Bisphenol S (BPS) is a chemical compound commonly used as a substitute for Bisphenol A (BPA) in various consumer products, particularly plastics, epoxy resins, and thermal paper (**Figure 2**).

**Figure 2:**
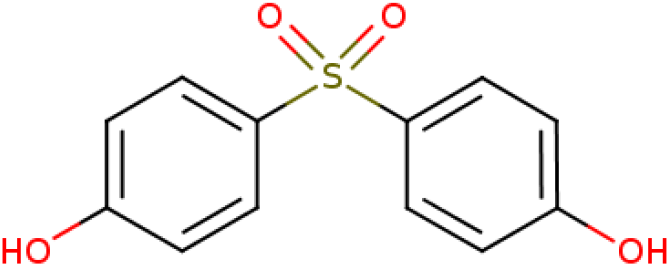
BPS or 4,4’-sulfonyldiphenol

While initially considered a safer alternative to BPA due to its lower estrogenic activity, BPS may still exert biological effects, particularly endocrinologically, according to emerging research. Studies have suggested that BPS can interact with hormone receptors, including estrogen receptors, albeit with lower potency compared to BPA. Like BPA, BPS has been linked to potential endocrine-disrupting consequences, impacting processes such as hormone signaling, reproduction, and development. Furthermore, BPS has been extensively detected in human biological samples, indicating widespread exposure. Concerns regarding its potential health impacts have led to increased scrutiny and calls for further investigation into its safety profile [11]. Research on the biological effects of BPS specifically on skeletal muscles is relatively limited compared to its repercussions on other systems like the endocrine and reproductive systems. However, one study showed that BPS reduces locomotor performance and modifies muscle protein levels, but not mitochondrial bioenergetics, in adult zebrafish [12].

## Bisphenol F

Bisphenol F (BPF) is another compound used as a substitute for BPA in various products (**Figure 3**). Similar to BPS, research on the specific biological effects of BPF on skeletal muscles is limited, but some general findings regarding its potential health impacts are known.

**Figure 3:**
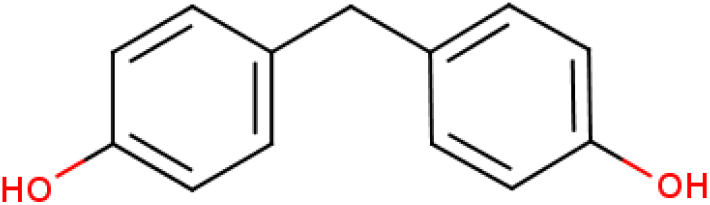
BPF or 4,4’-methylenediphenol

Like BPA and BPS, BPF has been shown to have endocrine-disrupting properties, although its potency may vary. Endocrine disruptors can interfere with hormone-signaling pathways, which are involved in regulating various physiological processes, including those related to skeletal muscle function and metabolism. While direct studies on BPF’s effects on skeletal muscles are scarce, research on other endocrine-disrupting compounds suggests that they can influence muscle health. For example, it has been shown that BPF induces motor degeneration and myelination defects in zebrafish larvae [13].

Overall, while more research is needed to elucidate fully the direct effects of BPS and BSF on skeletal muscles, existing evidence suggests that exposure to these compounds may have the potential to disrupt muscle function and physiology, either through direct effects on muscle cells or indirectly through endocrine disruption and metabolic disturbances. However, more research is needed to understand whether the roles of BPS and BPF are constructive or destructive.

## Discussion

FSHD2 is a complex genetic disorder characterized by progressive muscle weakness, primarily affecting facial, shoulder, and upper arm muscles. FSHD2 is caused by mutations in the SMCHD1 gene, leading to the aberrant activation of the DUX4 gene in adult muscle cells. Despite advancements, the pathophysiology of FSHD2 remains incompletely understood, and effective treatments are currently lacking.

In this study, we employed single-cell transcriptomic analysis to explore the molecular mechanisms underlying FSHD2. Such an approach offers a powerful means to dissect cellular heterogeneity and identify gene expression patterns at the individual cell level, offering valuable insights into disease pathogenesis. Our examination revealed significant dysregulation of genes essential for muscle function and integrity, shedding light on potential targets for therapeutic intervention. The upregulated genes identified in single-cell analysis encompass a diverse range of biological processes, including extracellular matrix regulation, cell growth, adhesion, and immune response. Notably, the upregulation of such genes as TIMP1, IGFBP7, and FN1 suggests aberrant tissue remodeling and inflammation, which may contribute to muscle degeneration in FSHD2. Furthermore, the downregulation of genes crucial for muscle contraction and sarcomere organization underscores the disruption of fundamental mechanisms governing muscle function.

Notably, our investigation unearthed the potential of BPS and BPF in modulating the expression of dysregulated genes in FSHD2. BPS exhibited a remarkable capacity to upregulate 52 of the top 100 downregulated genes in FSHD2, without concomitant downregulation of any other genes. Similarly, BPF demonstrated the ability to upregulate 45 of the top 100 downregulated genes, with only one gene undergoing downregulation. The enrichment analysis of these dysregulated genes underscored their profound involvement in diverse facets of muscle biology, as particularly pertaining to cardiac and skeletal muscle function. While our findings shed light on the potential therapeutic avenues involving BPS and BPF, acknowledging the nuanced role of these compounds in FSHD2 pathogenesis is imperative. Further research is warranted to delineate whether the modulation exerted by BPS and BPF is beneficial or detrimental in the context of FSHD2 progression.

## Summary

Our study offers novel insights into the molecular mechanisms driving FSHD2, highlighting potential therapeutic targets and environmental factors that may influence disease pathogenesis. By leveraging single-cell transcriptomics and gene-compound analysis, we have begun to unravel the complex interplay between genetic predisposition and environmental exposures in FSHD2, identified key dysregulated genes in FSHD2, and elucidated their modulation by compounds such as BPS and BPF. We are optimistic that this study has laid the groundwork for targeted interventions and personalized-medicine approaches in managing this debilitating disorder.

## Data Availability

https://www.ncbi.nlm.nih.gov/geo/query/acc.cgi?acc=GSE143452

